# Integrated Stepped-Care Task-Shared Intervention for Depression Symptoms in Older Adults Living with HIV in Nigeria: A pilot randomized controlled study

**DOI:** 10.64898/2026.11.22.26368694

**Authors:** Olufisayo Elugbadebo, Faithful Moposi Akintade, Tunde T. Omoloye, Bibilola D. Oladeji

## Abstract

**Background:** Depression can affect treatment adherence and quality of life of older adults living with HIV, yet evidence on integrated stepped-care task-shared (SCT) interventions in African settings remains limited. We assessed the feasibility of an adapted SCT intervention among adults aged ≥50 years receiving HIV care in Ibadan, Nigeria and examined preliminary differences in depressive symptoms, quality of life and ART adherence.

**Methods:** This was a two-arm, parallel-group, randomised pilot superiority trial embedded in a mixed-methods effectiveness-implementation study in Ibadan, Nigeria. Adults aged ≥50 years living with HIV and scoring ≥10 on the Patient Health Questionnaire-9 (PHQ-9) were randomised (1:1) to stepped-care task-sharing (SCT; n=50) or treatment as usual (TAU; n=50). SCT involved psychosocial sessions delivered by trained HIV clinic providers, with antidepressant treatment and specialist review according to depression severity and treatment response. TAU involved routine HIV care and referral to a separate psychiatric clinic. Outcomes were assessed at six weeks, three months and six months by assessors blinded to allocation. Feasibility indicators included recruitment, retention and intervention delivery. Preliminary clinical effectiveness was assessed using mixed-effects models for PHQ-9, quality of life (WHOQOL-HIV BREF) and ART adherence (Simplified Medication Adherence Questionnaire). Analyses followed randomised allocation using Stata/MP 15. The trial was registered with ClinicalTrials.gov (NCT06894680)

**Results:** Of the 757 participants screened, 120 participants were eligible and 100 (83.3%) were randomised, with 50 allocated to the SCT and 50 to TAU. All 50 SCT participants received the first psychosocial session, 40 (80%) attended at least two sessions and 30 (60%) completed all six sessions. Of 23 participants prescribed antidepressants, five (21.7%) initiated treatment. Baseline mean PHQ-9 scores were 13.24 (SD 4.43) in SCT and 14.12 (SD 4.48) in TAU. Of the 17 participants in TAU referred to the UCH psychiatric clinic, one attended but subsequently disengaged from care. Adjusted between-group differences (TAU minus SCT) favoured SCT at six weeks (3.95 points, 95% CI 1.91–5.99; p<0.001), three months (3.66, 95% CI 1.60–5.71; p<0.001) and six months (3.15, 95% CI 1.09–5.20; p=0.003). Quality-of-life scores increased in both groups, with no significant between-group differences at baseline, 3 months, or 6 months (all *p*>0.05). Differences in quality of life and ART adherence were not statistically significant. No adverse events were recorded.

**Conclusion:** This pilot trial demonstrated that the recruitment, retention and psychosocial SCT delivery were feasible within routine HIV care. SCT showed promising reductions in depressive symptoms over six months, although antidepressant initiation was limited. Further multisite evaluation should examine clinical effectiveness, intervention fidelity and implementation outcomes.

**Impact Statement:** Depression among older adults living with HIV can affect quality of life, treatment adherence, and overall well-being, yet mental health care is often not routinely integrated into HIV services, particularly in resource-constrained settings. This study provides evidence on the feasibility of integrating a stepped-care, task-shared approach to depression management into existing HIV care and treatment in Nigeria. By involving non-specialist healthcare providers in delivering psychosocial support and facilitating access to specialist care when needed, the intervention offers a potentially practical approach to addressing the mental health needs of older adults without relying exclusively on scarce mental health professionals. The findings may inform the development and implementation of more accessible, sustainable, and integrated mental health services for ageing populations living with HIV in Nigeria and other similar settings. Ultimately, this research contributes to efforts to ensure that people ageing with HIV receive comprehensive care that addresses not only their physical health but also their mental health and quality of life.

## Background

Depression is a common comorbidity among older adults (ages ≥50 years) living with HIV and can be associated with poor adherence to antiretroviral therapy (ART), reduced quality of life, adverse HIV-related outcomes, and increased mortality.^1–4^ Older adults living with HIV also experience a substantial burden of multimorbidity, which may complicate the management of depression alongside long-term HIV care.^5^ These challenges are concerning in sub-Saharan Africa (SSA), where the burden of HIV remains high and access to mental health services is limited.^6–9^ As more people living with HIV survive into later life, there is a growing need to integrate depression treatment into routine HIV treatment.^5,10^ Depression care that are specialist dependent are difficult to scale especially in resource-constrained settings where mental health professionals are scarce, and mental health care is often not an integral part of routine HIV services.^9,11^ For older adults already attending regular HIV appointments, referral to separate psychiatric services may involve additional visits, transport costs and difficulties maintaining treatment.^12,13^

Stepped-care and task-shared approaches offers an alternative by enabling trained non-specialist healthcare providers to deliver first line depression treatment, with treatment intensity determined by symptom severity and response to treatment and specialist care reserved for those requiring more intensive management.^11,14^ SCT combines psychological intervention, regular symptom monitoring, and treatment according to patient’s need and response to treatment^15^ . Evidence from collaborative and stepped-care trials supports improved depressive outcomes. The IMPACT trials reported improved depression outcomes among older adults receiving collaborative care, ^16^ while the CASPER trial demonstrated reductions in depressive symptoms among older adults with subthreshold depression.^17^

More recently, the HIV+D trial in Uganda showed that collaborative stepped-care integrated into routine HIV services improved depressive symptoms among adults living with HIV, with an adjusted PHQ-9 difference of 4.4 points (95% CI 3.4–5.5) at three months.^18^ However, IMPACT and CASPER involved older adults outside HIV treatment settings while participants in the Ugandan trial were younger, (mean age of 38 years).^18^ A systematic review by Mwangala et al. identified 50 studies involving people living with HIV aged ≥50 years in SSA, most of which were cross-sectional, with only one reporting data from a randomised trial.^19^ Evidence on the feasibility and clinical outcomes of integrated depression treatment in this population therefore remains limited.

In our earlier study of 400 participants, 25.8% met diagnostic criteria for major depressive disorder, of whom 79.6% had never received biomedical specialist care for depression^20^ Our subsequent pre-implementation study identified provider workload, limited training, stigma, discontinuity of care and financial constraints as barriers and leadership support and alignment with ART appointments as potential facilitators.^21^ These findings informed the adaptation of an SCT intervention combining low-intensity psychological treatment delivered by non-specialist providers, symptom monitoring, treatment intensification when indicated and specialist supervision, with flexible face-to-face or mobile phone contacts.

Although stepped-care interventions have demonstrated clinical benefits in other populations^22,23^, it remains important to establish whether this adapted model can be delivered within routine HIV services for older adults and whether it shows promising clinical benefit. We therefore conducted a pilot randomized controlled trial among adults aged ≥50 years receiving HIV care in Ibadan, Nigeria. The study examined the feasibility of recruitment, retention and SCT delivery, and estimated between-group differences in depressive symptoms at six weeks, three months and six months. We also explored changes in quality of life and ART adherence. Given the pilot design, the clinical findings were intended to inform a future fully powered effectiveness trial.

## Methods

### Trial design

This pilot two-arm, parallel group randomised controlled superiority trial (registered at Clinical Trials.gov, trial registration number-NCT06894680) evaluated the preliminary effectiveness of an adapted stepped-care task-sharing (SCT) intervention for depression among adults aged ≥ 50 years living with HIV receiving care at a large HIV clinic in Ibadan. Participants were randomly assigned in a 1:1 ratio to either the SCT intervention or treatment as usual (TAU) using a computerized randomisation sequence, with 50 participants allocated to each group. The trial was embedded within a broader mixed-methods effectiveness-implementation study that examined contextual factors influencing the integration of depression care and the findings were used to inform adaptation of the SCT intervention.

### Trial Setting

This study was conducted at the HIV clinic of the Infectious Disease Institute (IDI), University of Ibadan/ University College Hospital, Nigeria. It serves as a major referral centre providing care to approximately 6,540 people living with HIV as of June 2026. Adults aged 50 years and older constituted about 53% of the clinic population. Routine services include clinical care, counselling, laboratory diagnostics, pharmacy and medication pick-up, and follow-up, delivered by a multidisciplinary team of physicians, nurses, pharmacists, counsellors, laboratory scientists, and records officers. Routine mental health screening was not part of standard HIV care. Patients with identified mental health problems received counselling and medical officer review, with referral for specialist care to the UCH psychiatric clinic at a separate location when indicated.

### Patient and public involvement

The Patients and Health Care Providers were consulted before the study commenced, and their input informed the design and planned delivery of the intervention.

### Participants and recruitment

Participants were adults aged ≥50 years living with HIV and receiving care at the study clinic. During the recruitment period, research staff approached clinic attendees, provided information about the study, and were screened for depressive symptoms using the 9-item Patient Health Questionnaire (PHQ-9)^26^. Individuals ≥50 years were eligible if they had a PHQ-9 score of ≥10 and provided informed consent to participate. A PHQ-9 score of ≥10 was selected as the eligibility threshold because it indicates at least moderate depressive symptom severity and has been previously used in HIV care trials ^18^ ^27^. Participants were excluded if they were receiving treatment for depression or any other previously diagnosed mental disorder, required urgent psychiatric assessment, were at imminent risk of suicide, had severe cognitive impairment that could compromise informed consent or study participation, or had comorbidities that contraindicated sertraline use. Eligible participants who provided informed consent completed baseline assessment before randomization. Participants were randomly allocated in a 1:1 ratio to either the SCT or TAU group using a computer-generated randomisation sequence. (see section on randomization below).

### Intervention and Comparator

#### Stepped-care task-sharing (SCT) intervention

This trial adopted an SCT intervention for the integration of depression treatment into the HIV clinic. Treatment intensity was determined by baseline depression severity and response to treatment. Participants with baseline PHQ-9 of 10-14 commenced step 1 (psychosocial intervention) while those with scores ≥15 commenced Step 2 (psychosocial intervention plus antidepressant; sertraline prescribed by the clinic physician) (Fig 2). Trained clinic healthcare providers delivered the psychosocial intervention, monitored participants during treatment, ensured participants requiring medications were reviewed by the clinic physicians and facilitated physician or psychiatrist review when indicated.

At 6 weeks, trained research assistants administered the PHQ-9 and provided scores to the intervention provider. Participants with persistent or worsening symptoms (PHQ-9 ≥10) following the first six weeks were stepped up: those in Step 1 received additional sertraline treatment, starting at 50 mg daily, while those in Step 2 received Step 3 specialist psychiatric review for individualised medication adjustment and/or increased psychosocial sessions. Clinic physicians and intervention providers implemented the psychiatrist’s treatment plan.

The psychosocial intervention adapted from the WHO Mental Health Gap Action Programme Intervention Guide (mhGAP-IG)^28^ depression module included psychoeducation, behavioural activation, problem-solving, stress management, and strengthening social support. The providers received a 3-day training before delivering the intervention. The psychosocial intervention was conducted weekly, each lasting approximately 30 - 45 minutes, over the initial 6-week treatment period. Sessions were scheduled in a flexible manner as recommended during the pre-implementation phase^21^ and were delivered either face-to-face or by mobile phone. Where possible, sessions were aligned with ART pickup. Heath care providers monitored participants weekly, using a checklist adapted from the Problem Management Plus guide^28^ and received supportive supervision from a psychiatrist fortnightly to address clinical concerns and delivery challenges. Fidelity was monitored using a checklist covering the core components of the psychosocial intervention and adherence to treatment procedures at each level of care.

### Treatment as Usual (TAU)

Participants allocated to TAU continued to receive routine HIV care at the study clinic. Those identified with depression were referred to the psychiatry clinic at the University College Hospital (UCH), Ibadan, for further assessment and treatment in accordance with the existing care practices. TAU participants did not receive the structured SCT intervention within the HIV clinic. Participants were followed up via the mobile phone to determine referral attendance, treatment received and any additional mental health care accessed elsewhere.

### Stepped-Care Treatment (SCT) Pathway

Delivered primarily by trained non-specialist HIV care providers under structured specialist supervision

### Treatment as usual

Referrals were formally documented but the study team did not facilitate appointments at the psychiatric centre.

### Outcome measures and assessment schedule

#### Outcome assessment

Outcome assessments were conducted at baseline and the prespecified follow-up time points. Trained research assistants blinded to treatment allocation administered the outcome measures at the scheduled assessment points. Feasibility indicators were assessed separately using intervention records and treatment documentation.

### Feasibility outcomes

#### Feasibility indicators for the Intervention

Feasibility was assessed using intervention delivery, treatment uptake, and referral. These included the proportions of participants who received at least one, two, or all planned psychosocial intervention sessions; physician contact, antidepressant initiation and completion and psychiatric consultation when indicated. Data were obtained from intervention and treatment records using eligible participants as the denominator. The adopted prespecified benchmarks were similar to those used in a pilot trial of a SCT intervention package for depression in primary healthcare conducted in Nigeria^22^. ^29^.

#### Preliminary Clinical Effectiveness Outcome

Depressive symptom severity, measured using the PHQ-9, was the registered primary clinical outcome, assessed at baseline, six weeks, three months and six months. Between group differences were estimated at each follow-up to examine the magnitude and direction of potential treatment effects. Given the pilot design, these analyses were exploratory, and effect estimates with 95% confidence intervals were interpreted as preliminary findings to inform the design and parameters needed for a future fully powered effectiveness trial. Secondary outcomes included depression remission, quality of life and ART adherence.

#### Secondary Outcomes

Secondary outcomes were quality of life and ART adherence. Quality of Life was assessed using WHOQOL-HIV BREF, a 31-item instrument developed by the World Health Organization to assess quality of life among people living with HIV^30^. The instrument measures six domains: physical, psychological, level of independence, social relationships, environment, and spirituality/religion/personal beliefs. Items are rated on a 5-point response scale, with higher scores indicating better quality of life. The WHOQOL-HIV BREF was administered at baseline, 3 months, and 6 months. Medication adherence was assessed using the Simplified Medication Adherence Questionnaire (SMAQ), a six-item self-report measure developed to assess adherence to antiretroviral therapy^31^The questionnaire assesses missed doses, difficulties with medication-taking and patterns of non-adherence. Participants were classified as adherent or non-adherent using established SMAQ scoring criteria, Assessment were conducted at baseline, 3 months, and 6 months.

#### Implementation Outcomes

Acceptability, appropriateness, and perceived feasibility of intervention were assessed at baseline and post-intervention using the Acceptability of Intervention Measure (AIM), Intervention Appropriateness Measure (IAM), and Feasibility of Intervention Measure (FIM), respectively. Each measure comprises four items, giving a total of 12 items across the three measures. Responses were recorded on a five-point Likert scale, with higher scores indicating greater acceptability, appropriateness, and perceived feasibility, respectively.^32^

### Sample size

The sample size was calculated for a two-arm randomised controlled trial designed to evaluate the superiority of the intervention over treatment as usual. The calculation used a Z-score of 1.96 for the significance level and 0.84 for statistical power. The estimated sample size was subsequently adjusted for an anticipated attrition rate of 25% to account for potential loss to follow-up. Following this adjustment, the target sample size was 50 participants per group, giving a total sample of 100 participants, with 50 participants allocated to the intervention group and 50 to the control group. As a pilot trial, the study was not designed to provide definitive evidence of treatment effectiveness; rather, the sample was intended to provide preliminary estimates of treatment effects and inform a subsequent fully powered trial.

### Randomisation, allocation concealment, and masking

Following screening and baseline assessment, participants were randomly allocated in a 1:1 ratio to the SCT intervention or TAU. The study statistician generated the randomisation sequence using a computer-generated sequence. Allocation was concealed using sequentially numbered, opaque, sealed envelopes, which were opened after completion of the baseline assessment. Trained research assistants enrolled participants, while a designated member of the research team (FM) implemented the treatment allocation. Given the nature of the intervention, care providers were aware of the treatment they delivered. Outcome assessors were blinded to treatment allocation and were not involved in participant screening or recruitment

## Statistical analysis

Analyses were conducted using Stata version MP 15. Baseline characteristics were summarised by randomised treatment group. Feasibility outcomes were reported as frequencies and percentages using appropriate denominators. Participants were analysed according to their randomised allocation regardless of intervention completion.

Preliminary clinical effectiveness was assessed using PHQ-9 scores at baseline, 6 weeks, 3 months and 6 months. A linear mixed-effects model included the treatment group, assessment time, and their interaction as fixed effects, with a participant level random intercept to account for repeated measurements. Adjusted mean PHQ-9 scores and between-group differences were reported with 95% confidence intervals and Cohen’s d. The six-month between-group difference was examined as an exploratory estimate of treatment effect. Remission (PHQ-9 <5) was analysed using separate binary logistic regression models at each follow up, reporting odds ratios and 95% confidence intervals. BREF scores and SMAQ adherence at baseline, three and six months were analysed using linear and logistic mixed-effects models, respectively. Adjusted mean differences and predicted adherence probabilities were reported with 95% confidence intervals. Model assumptions for continuous outcomes were examined using residual plots

### Missing data and sensitivity analyses

Follow-up completion and reasons for missing assessments were recorded at 6 weeks, 3 months, and 6 months. Patterns of missing data were examined by treatment group. Linear mixed-effects analyses included all available repeated measurements under the missing-at-random assumption. Participants who discontinued the intervention but completed follow-up assessments were retained in their original treatment groups. No multiple imputation or per-protocol sensitivity analyses were performed. Remission analyses included participants with available PHQ-9 data at each assessment.

### Ethical approval and trial registration

Ethical approval was obtained from the University of Ibadan/University College Hospital (UI/UCH) Ethics Committee with IRB number (UI/EC/24/01020). All participants provided written informed consent before enrolment. The trial was registered with ClinicalTrials.gov (registration number NCT06894680; registration date (03/18/2025).

## RESULTS

### Participants flow, Recruitment, intervention delivery and follow-up

Between March and May 2025, a total of 757 older people living with HIV were screened for eligibility, of whom 120 (15.9%) were eligible. One declined participation, while 19 refused participated because they could not commit to the follow up process. The remaining 100 participants (83.3% of those eligible) were randomized equally to SCT and TAU, giving a recruitment yield of 13.2%. At six months, follow-up assessments were completed by 46 participants (92%) in each group (Figure 1). Most participants lost to follow-up could not be reached despite at least four telephone contact attempts and had also missed their scheduled antiretroviral medication pick-up visits. One participant died during follow-up. No demographic or clinical features were associated with refusal to participate or with attrition. (Figure 2)

**Fig 1.**
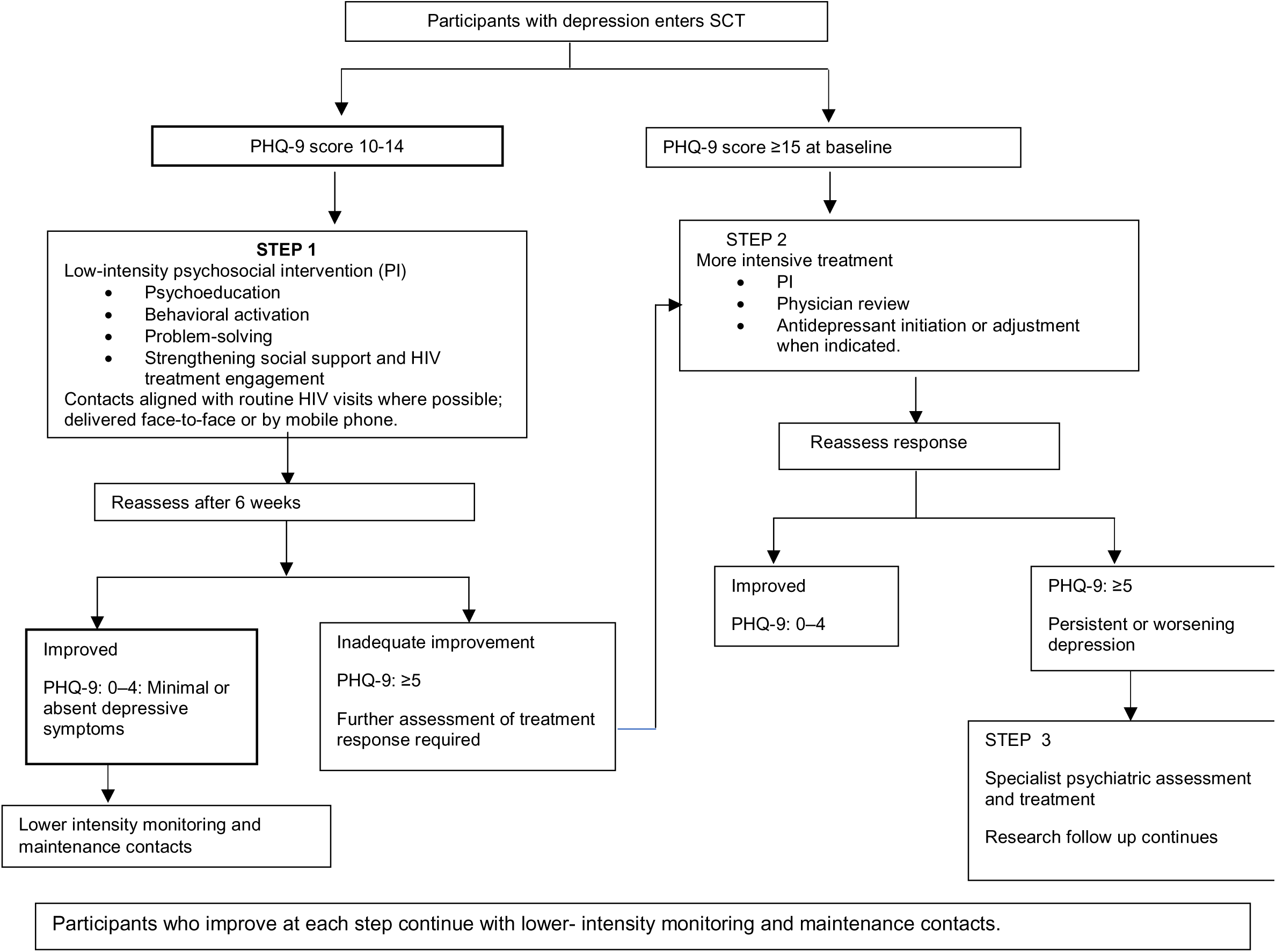
Intervention Flowchart.

**Figure 2:**
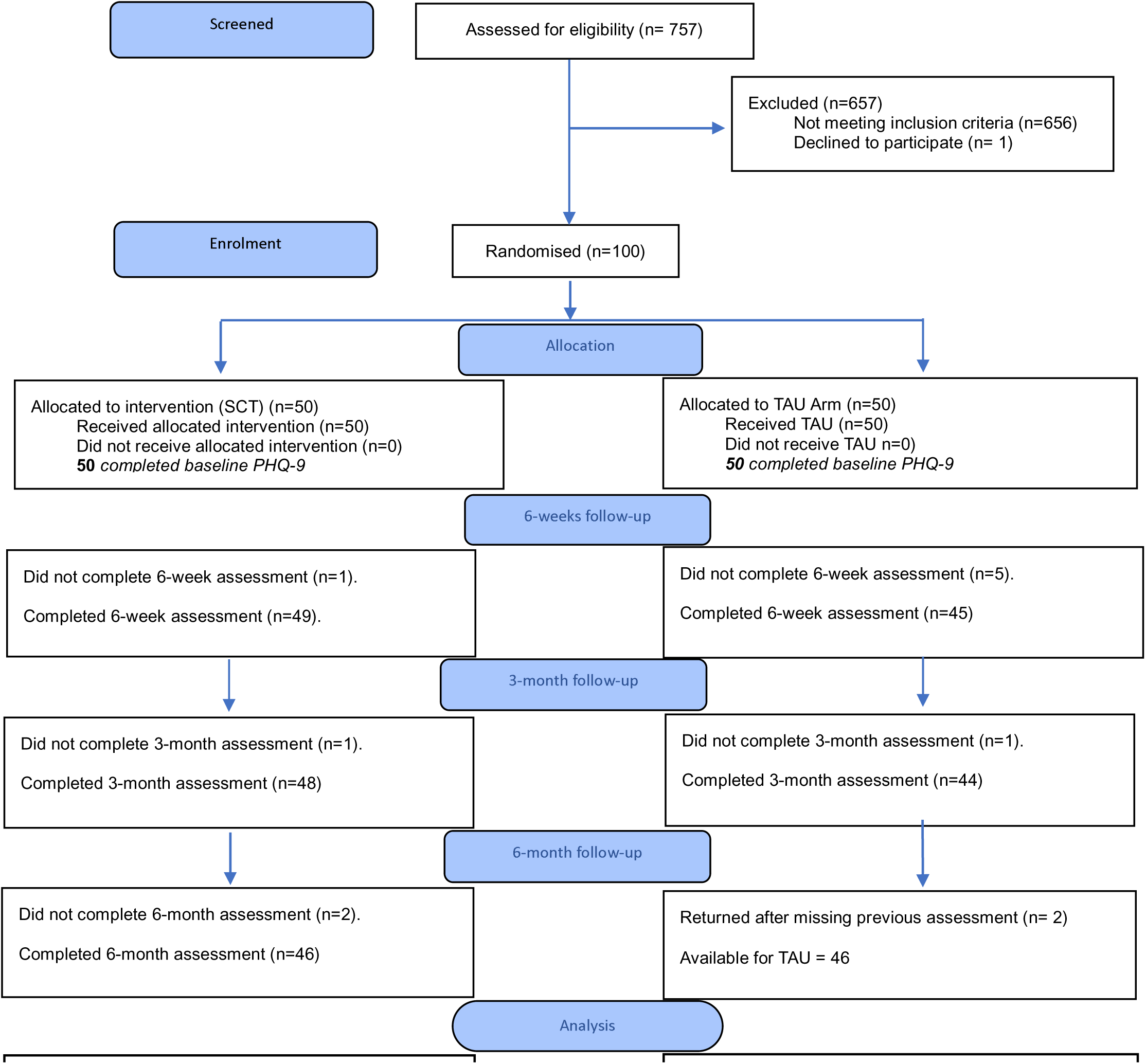
CONSORT participant flow. Patient Health Questionnaire-9 (PHQ-9) for assessing the severity of depressive symptoms over the preceding two weeks.

### Process indicators for the intervention arm

All 50 participants (100%) received the first psychosocial intervention session, exceeding the 95% benchmark. Physician contact was recorded for 16 participants (32%), while 13 (26%) received psychiatric consultation. Antidepressants were prescribed to 23 participants (46%), of whom five (21.7%) initiated treatment. Among the 18 who did not initiate treatment, 15 (83.3%) cited financial constraints and reported that they would have taken sertraline if provided free of charge, while three (16.7%) did not perceive a need for antidepressants. (Table 1)

**Table 1:** Feasibility indicators for intervention delivery.

| **Process indicators** | **Number** | **Percentage (%)** | **Projected benchmark (%)** |
|---|---|---|---|
| Proportion of participants who received the first PI session | 50 | 100 | 95 |
| Proportion who received at least 2 PI sessions | 40/50 | 80 | 70 |
| Proportion who completed the first 6 weeks PI intervention | 30/50 | 60 | 50 |
| **Treatment uptake and referral** |  |  |  |
| Proportion with Physician contact | 16/50 | 32 |  |
| Proportion who received psychiatrist consultation | 13/50 | 26 |  |
| Proportion who received antidepressant prescription | 23/50 | 46 |  |
| Proportion who initiated antidepressant | 5/23 | 21.7 | 20 |

### Referral uptake in the TAU arm

Participants in the TAU arm who required additional mental health care were referred to the UCH psychiatric clinic. Of the 17 participants referred, one attended but subsequently disengaged from care.

### Acceptability, Appropriateness and Perceived Feasibility

Among the 11 patients and five healthcare providers assessed, mean acceptability scores were 4.55 and 4.65, respectively. Corresponding scores for appropriateness were 4.55 in both groups, while perceived feasibility scores were 4.50 and 4.60, respectively.

### Baseline Characteristics of the Participants

At baseline, the Intervention and TAU groups had similar mean ages; 58.35 years (SD = 7.70) in the Intervention group compared to 57.96 years (SD = 7.48) in the TAU group. There were more women accounting for 70.0% of the participants in the intervention group and 82.0% of the TAU group. Viral load status was identical across both treatment groups with similar proportions (94%) virally suppressed (viral load; ≤1,000 copies/mL at all available time points), unsustained viral suppression (any viremia of at least one measurement >1,000 copies/mL during the study period). The baseline PHQ-9 scores were also comparable, mean scores was 13.24 (SD = 4.43) in the Intervention group and 14.12 (SD = 4.48) in the TAU group. (Table 2)

**Table 2:** Baseline characteristics of participants.

| Characteristics | Intervention (n = 50),<br>n (%) | TAU (n = 50),<br>n (%) |
|---|---|---|
| **Age category** |  |  |
| 50–64 years | 31 (62.0) | 36 (72.0) |
| 65–74 years | 5 (10.0) | 8 (16.0) |
| 75–85 years | 1 (2.0) | 3 (6.0) |
| Missing | 13 (26.0) | 3 (6.0) |
| **Gender** |  |  |
| Male | 13 (26.0) | 9 (18.0) |
| Female | 35 (70.0) | 41 (82.0) |
| Missing | 2 (4.0) | 0 (0.0) |
| **Educational attainment** |  |  |
| No formal education | 10 (20.0) | 15 (30.0) |
| Primary | 9 (18.0) | 7 (14.0) |
| Secondary | 8 (16.0) | 11 (22.0) |
| Tertiary | 16 (32.0) | 17 (34.0) |
| Missing | 7 (14.0) | 0 (0.0) |
| **Marital status** |  |  |
| Single | 1 (2.0) | 5 (10.0) |
| Married | 17 (34.0) | 17 (34.0) |
| Widowed | 23 (46.0) | 27 (54.0) |
| Divorced | 2 (4.0) | 1 (2.0) |
| Missing | 7 (14.0) | 0 (0.0) |
| **Viral load status** |  |  |
| Suppressed | 47 (94.0) | 47 (94.0) |
| Non-suppressed | 3 (6.0) | 3 (6.0) |
|  | **Mean (SD)** | **Mean (SD)** |
| **PHQ-9 total score** | 13.24 (4.43) | 14.12 (4.48) |
| **SMAQ** | 0.64(0.48) | 0.60(0.49) |
| **WHOQOL-BREF** | 95.62 (13.16) | 97.53 (13.21) |

### Preliminary clinical outcome: depressive symptoms over 6 months

The mixed-effects analysis included 378 PHQ-9 observations from 100 randomised participants. Depressive symptom trajectories differed significantly between SCT and TAU over six months (treatment group × time: Wald χ²(3) = 9.64, *p* = .022). Although PHQ-9 scores decreased in both groups, reductions were greater in SCT. The adjusted between-group differences (TAU − SCT) were 3.95 points at 6 weeks (Cohen’s d = −0.71), 3.66 at 3 months (d = −0.66) and 3.15 at 6 months (d = −0.60), indicating moderate to large effects favouring SCT, although the magnitude decreased over time (Table 3).

**Table 3.**
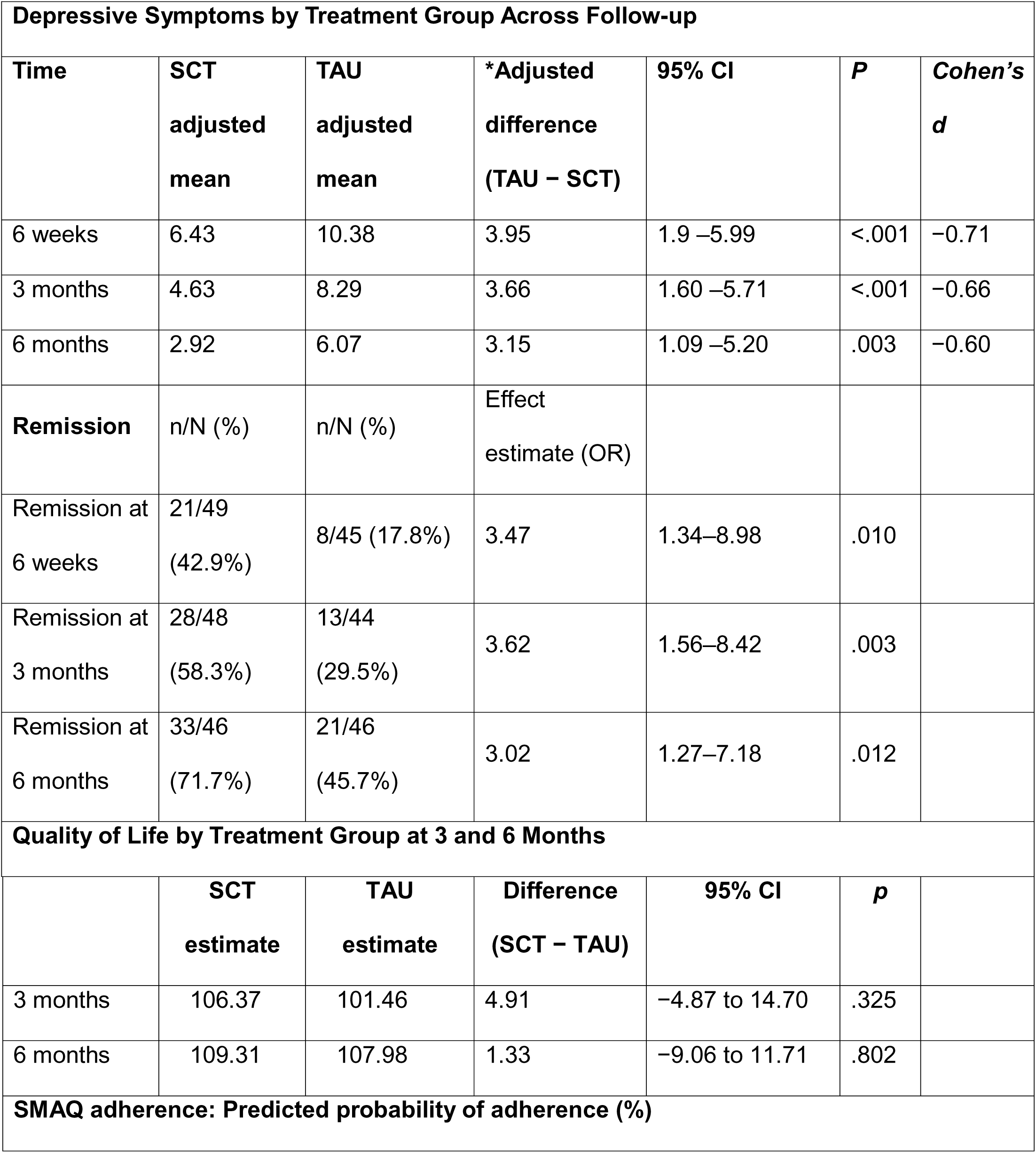
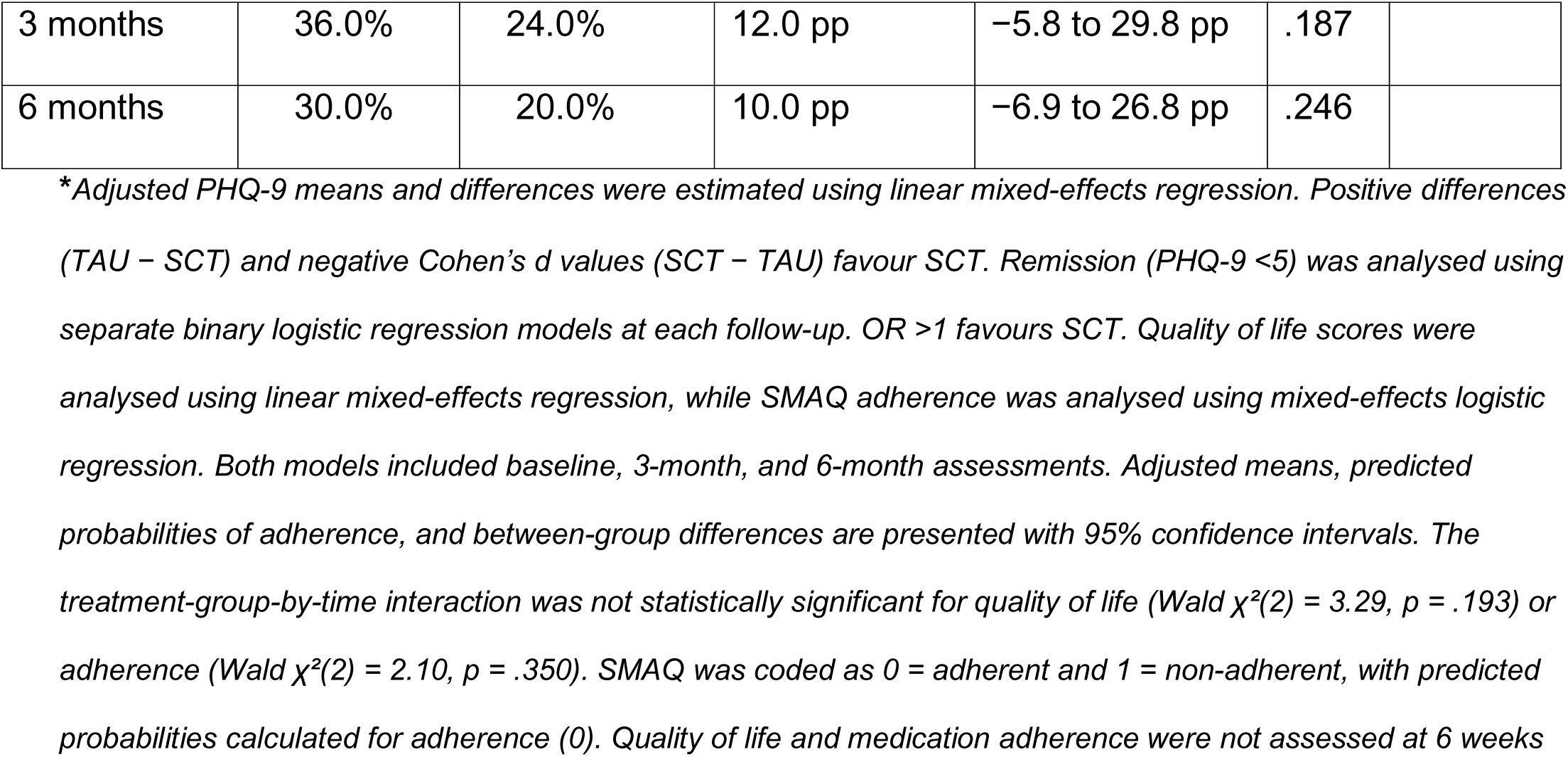
Depressive Symptoms, Remission, Quality of Life and Antiretroviral Medication Adherence by Treatment Group Across Follow-up.

| **Depressive Symptoms by Treatment Group Across Follow-up** |  |  |  |  |  |  |
|---|---|---|---|---|---|---|
| **Time** | **SCT adjusted mean** | **TAU adjusted mean** | **\*Adjusted difference (TAU − SCT)** | **95% CI** | ***P*** | ***Cohen’s d*** |
| 6 weeks | 6.43 | 10.38 | 3.95 | 1.9 –5.99 | <.001 | −0.71 |
| 3 months | 4.63 | 8.29 | 3.66 | 1.60 –5.71 | <.001 | −0.66 |
| 6 months | 2.92 | 6.07 | 3.15 | 1.09 –5.20 | .003 | −0.60 |
| **Remission** | n/N (%) | n/N (%) | Effect estimate (OR) |  |  |  |
| Remission at 6 weeks | 21/49 (42.9%) | 8/45 (17.8%) | 3.47 | 1.34–8.98 | .010 |  |
| Remission at 3 months | 28/48 (58.3%) | 13/44 (29.5%) | 3.62 | 1.56–8.42 | .003 |  |
| Remission at 6 months | 33/46 (71.7%) | 21/46 (45.7%) | 3.02 | 1.27–7.18 | .012 |  |
| **Quality of Life by Treatment Group at 3 and 6 Months** |  |  |  |  |  |  |
|  | **SCT estimate** | **TAU estimate** | **Difference (SCT − TAU)** | **95% CI** | ***p*** |  |
| 3 months | 106.37 | 101.46 | 4.91 | −4.87 to 14.70 | .325 |  |
| 6 months | 109.31 | 107.98 | 1.33 | −9.06 to 11.71 | .802 |  |
| **SMAQ adherence: Predicted probability of adherence (%)** |  |  |  |  |  |  |
| 3 months | 36.0% | 24.0% | 12.0 pp | −5.8 to 29.8 pp | .187 |  |
| 6 months | 30.0% | 20.0% | 10.0 pp | −6.9 to 26.8 pp | .246 |  |

\**Adjusted PHQ-9 means and differences were estimated using linear mixed-effects regression. Positive differences (TAU − SCT) and negative Cohen’s d values (SCT − TAU) favour SCT. Remission (PHQ-9 <5) was analysed using separate binary logistic regression models at each follow-up. OR >1 favours SCT. Quality of life scores were analysed using linear mixed-effects regression, while SMAQ adherence was analysed using mixed-effects logistic regression. Both models included baseline, 3-month, and 6-month assessments. Adjusted means, predicted probabilities of adherence, and between-group differences are presented with 95% confidence intervals. The treatment-group-by-time interaction was not statistically significant for quality of life (Wald χ²(2) = 3.29, p = .193) or adherence (Wald χ²(2) = 2.10, p = .350). SMAQ was coded as 0 = adherent and 1 = non-adherent, with predicted probabilities calculated for adherence (0). Quality of life and medication adherence were not assessed at 6 weeks*

### Secondary Outcomes: Quality of life and Medication adherence

Quality of life scores increased in both groups, but the treatment-group-by-time interaction was not statistically significant (Wald χ²(2) = 3.29, *p* = .193). Although adjusted mean scores were higher in the intervention group at 3 and 6 months, the between-group differences were not statistically significant.

Medication adherence was slightly higher in TAU at baseline and in the intervention group at 3 and 6 months. However, neither the treatment-group-by-time interaction (Wald χ²(2) = 2.10, *p* = .350) nor the between-group differences at either follow-up were statistically significant (Table 3).

### Harms

No adverse events were recorded in this study. One participant died during the study, however, the death was unrelated to the study procedures.

## Discussion

In this pilot randomised controlled trial, we examined the feasibility of recruiting older adults living with HIV and delivering stepped-care task-sharing (SCT) for depression within routine HIV services. Of the 120 eligible participants, 100 (83.3%) were randomised, with 92% completing the six-month follow-up in both groups. All 50 participants allocated to SCT received the first psychosocial intervention session, 40 (80%) received at least two sessions, and 30 (60%) completed all six initial sessions. These proportions exceeded the reference benchmarks adapted from Oladeji et al. (2015), suggesting that psychosocial treatment delivered by non-specialist healthcare providers within routine HIV services are acceptable to the participants. The high follow-up completion also supports the feasibility of retaining older adults receiving HIV care in a trial involving repeated assessments over six months.

Our findings are consistent with findings from a similar study on SCT by Oladeji et al. (2015), who demonstrated the feasibility of delivering stepped-care depression treatment by primary healthcare workers in Nigeria, although sustaining attendance at psychological treatment sessions presented challenges^33^. In our study, face-to-face sessions were aligned with routine antiretroviral medication pick-up visits, while telephone sessions were used when appointments did not align with the visit. These adaptations were informed by our pre-implementation study, in which patients and healthcare providers identified transport costs, frequent clinic visits and limited provider capacity as potential barriers to intervention delivery.^21^ The favourable post intervention ratings of acceptability, appropriateness and perceived feasibility among patients and healthcare providers further support the practical delivery of SCT within the HIV clinic.

In this study, we observed that the uptake of antidepressant was lower than psychosocial intervention uptake. Of the 23 participants prescribed sertraline, five (21.7%) initiated treatment. Among the 18 who did not commence medication, 15 reported financial constraints and indicated that they would have taken the medication if it had been provided free of charge, while three did not perceive a need for antidepressants. This suggests that affordability, rather than unwillingness to receive antidepressant treatment, was the main reported barrier to medication initiation. Our pre-implementation study had already identified financial constraints and lack of antidepressant availability within the HIV clinic as barriers to integrated depression care. ^21^ In our setting, antiretroviral medication is provided free of charge, while patients generally have to pay out of pocket for antidepressant. Though ART medications are provided free of charge patients incurred costs for other healthcare service including mental healthcare among people receiving HIV treatment in Nigeria.^12,34^ These findings indicate that providing depression care within HIV services must also address access to prescribed medication.

The preliminary clinical findings favoured SCT, with greater reductions in depressive symptoms than TAU at six weeks, three months and six months. At three months, the adjusted between-group difference was 3.66 PHQ-9 points (Cohen’s d = −0.66), with a difference of 3.15 points maintained at six months (d = −0.60). This pattern is similar to findings from the Ugandan HIV+D trial, where collaborative stepped care produced greater reductions in depressive symptoms at three months, with benefits maintained, although attenuated, at 12 months.^18^ An important observation in our study was that improvement in depressive symptoms occurred despite limited initiation of prescribed antidepressants. This suggests that the psychosocial intervention, regular symptom monitoring and other components of structured depression care may be effective in reducing symptoms.

The greater reduction in depressive symptoms among SCT participants is also consistent with findings from the IMPACT trial, which demonstrated sustained improvements with collaborative care among older adults^16^ and the PROACTIVE trial in Brazil, which reported improved depression outcomes following a task-shared psychosocial intervention among older adults.^35^ Similarly, Safren et al. (2021) reported improvements in depression following nurse-delivered cognitive behavioural therapy integrated into HIV care in South Africa.^36^ However, these studies did not specifically focus on older adults receiving HIV treatment. Our study adds preliminary evidence on the feasibility and potential clinical benefits of integrated depression care in this population.

The nature of the comparison condition is important when interpreting our findings. While SCT participants received structured depression care within the HIV clinic, TAU participants were referred to a psychiatric clinic outside the HIV service. The greater reduction in depressive symptoms among SCT participants may therefore reflect both the intervention and improved access to depression treatment. However, our study could not differentiate the contribution of these factors.

Quality of life and ART adherence did not differ significantly between groups over the follow-up period. Although the estimated probability of ART adherence was higher in SCT at three and six months, the differences were not statistically significant. This differs from findings by Safren et al. (2021), whose intervention specifically addressed both depression and ART adherence. Our intervention focused primarily on depression, which may partly explain the absence of a clear difference in adherence. In addition, this pilot and was not powered to enable the exploration of treatment effects on these secondary outcomes.

## Limitations

This single-site pilot trial involved 100 participants, limiting generalisability. Despite high follow-up completion, missing outcome data may have introduced bias. As SCT combined psychosocial treatment, symptom monitoring, antidepressant medication and specialist review, the contribution of individual components could not be determined.

## Conclusion

This pilot trial demonstrated the feasibility of recruiting and retaining older adults living with HIV and delivering psychosocial depression care within routine HIV services. SCT was associated with greater reductions in depressive symptoms than treatment as usual, with the between-group difference maintained over six months. The main difficulty identified was access to prescribed antidepressant treatment, a barrier previously recognised during pre-implementation and further supported with this pilot trial. Findings support further evaluation of SCT in a larger multisite trial examining clinical effectiveness, intervention fidelity, implementation outcomes and resources required for sustained delivery.

## Data Availability

All data produced in the present study are available upon reasonable request to the authors

## Open Science

### Trial registration

The trial was registered with ClinicalTrials.gov (registration number NCT06894680; registration date (03/18/2025). Trial results have not been publicly posted on the registry, been published as a preprint or been published in a journal.

### Protocol and statistical analysis plan

The trial protocol is available at https://clinicaltrials.gov/study/NCT06894680.

### Data sharing

De-identified participants’ data, the data dictionary, and statistical code may be made available by the corresponding author upon reasonable request, subject to institutional approval and applicable ethical and data-protection requirements. Requests for intervention materials will be considered separately.

### Funding

This study was supported by the Royal Society of Tropical Medicine and Hygiene (RSTMH) (nihr24156). The funder had no role in the study design, data collection, data analysis, interpretation of findings, or preparation of the manuscript.

---

**Supplementary Table S1.** Mixed-Effects Model of PHQ-9 Scores by Treatment Group and Time.

| **Fixed effect** | **B** | **SE** | ***z*** | ***p*** | **95% CI** |
|---|---|---|---|---|---|
| **Group** |  |  |  |  |  |
| TAU vs SCT | 0.88 | 1.02 | 0.87 | .387 | −1.11 to 2.87 |
| **Time** |  |  |  |  |  |
| 6 weeks vs baseline | −6.81 | 0.77 | −8.83 | <.001 | −8.32 to −5.30 |
| 3 months vs baseline | −8.61 | 0.78 | −11.09 | <.001 | −10.13 to −7.09 |
| 6 months vs baseline | −10.32 | 0.79 | −13.11 | <.001 | −11.86 to −8.77 |
| **Group × time** |  |  |  |  |  |
| TAU × 6 weeks | 3.07 | 1.11 | 2.78 | .005 | 0.90 to 5.24 |
| TAU × 3 months | 2.78 | 1.11 | 2.49 | .013 | 0.59 to 4.96 |
| TAU × 6 months | 2.27 | 1.11 | 2.04 | .042 | 0.08 to 4.45 |

Wald χ²(3)=9.64, *p*=0.022.

